# Influence of Anemia on Prevalence of Gestational Diabetes among Pregnant Women in Tripoli, Libya

**DOI:** 10.1101/2024.04.07.24305457

**Authors:** Ahmed Atia, Hosam Elmahmoudi

## Abstract

Gestational diabetes mellitus (GDM) is raised globally leading to substantial maternal and foetal morbidity. This study aimed to determine the prevalence of GDM among pregnant women delivering in different private polyclinics in Tripoli, Libya. A cross-sectional study was conducted among pregnant women who were admitted to gynecology department of in different medical polyclinics, Tripoli, Libya from Jan to Dec 2022. The prevalence of GDM in pregnant women increased with increase of the gestational age to reach maximum (86%) at the third trimester of gestation. About 31% (96 patients) anemic pregnant women were observed in 312 GDM. Careful surveillance is required for these pregnancies in high-risk units for early detection and treatment of possible complications, in order to try to reduce maternal and neonatal morbidities.

## Introduction

Gestational diabetes (GDM) is defined as diabetes that develops for the first-time during pregnancy. GDM, like other types of diabetes, changes how your cells use sugar (glucose). Gestational diabetes is characterized by high blood sugar levels, which can have an impact on both pregnancy and baby health. It can occur at any stage of pregnancy, but is most common during the second or third trimester [1]. All pregnant women should be tested for gestational diabetes at 24-28 weeks of pregnancy (except those women who already have diabetes) [2].

GDM is one of the most common health problems for pregnant women. It affects about 5 percent of all pregnancies, which means there are about 200,000 cases each year. If not treated, gestational diabetes can cause health problems for mother and fetus [1]. It is crucial to comprehend the healthcare needs of a given population at a given time, and prevalence estimates are perfect for this [3]. Regrettably, ethnicity, ethnic variation within and between populations, and uneven application of screening and diagnostic criteria account for a large portion of the variation observed in the global estimates of GDM prevalence (<1%–28%) [4]. Ascertaining the region-specific prevalence estimate is crucial to accurately projecting the GDM burden of a given area. The Middle East and North Africa (MENA) region has a low literature on the prevalence of GDM, despite the fact that two major risk factors—physical inactivity and an above-normal body mass index (BMI)—are found to be highly prevalent there [5,6].

In Africa, A meta-analysis estimated that the overall prevalence of gestational diabetes in sub-Saharan Africa was 9% (95%CI, 7-12%) based on the pooling of data from 33 studies [7]. Only six nations in this part of the world have had their prevalence evaluated [8]. Moreover, not much research has been done on the GDM risk factors in black sub-Saharan African populations. As a result, in sub-Saharan Africa, traditional risk factors like age, being overweight, having a history of gestational diabetes, etc., are regarded as risk factors for GDM [9].

Anemia is the most common disorder in the world and is believed to cause a number of short- and long-term complications [10]. It also conveyed to lower the risk of the pregnant woman developing GDM [11]. However, recent genetic study reported elevated iron stores and increased GDM incidence in women with heterozygous hemoglobinopathies [12].

There is a dearth of information regarding the state of GDM in Libya. Additionally, data are not available, according to the IDF atlas [10]. There are no articles on GDM in the Libyan Medical Journal that discuss the condition’s prevalence, treatment, and aftercare. Therefore, the current study was conducted to assess the prevalence of GDM among pregnant women in Tripoli, Libya.

## Methods

### Study settings

A cross-sectional study design was used to assess the prevalence of GDM and its risk factors among pregnant women who were admitted to gynecology department of in different medical polyclinics, Tripoli, Libya from Jun to Dec 2022.

### Data collection

Women who visited the gynecology department in different medical polyclinics, Tripoli, Libya during the study period participated in the study. After obtaining written informed consent, two milliliters of venous blood were collected under aseptic conditions and transferred in to glucose tubes by mixing for 5 minutes. The specimens were labeled with identification number of each study participant. The blood sugar test was determined using Cobas Integra 400 Plus which applies fully automated analyzer.

The reliability of the study findings was guaranteed by implementing quality control (QC) measures throughout the whole process of the laboratory work. All materials, equipment and procedures were adequately controlled. Appropriate volume of blood and anticoagulant was used to maintain the specimen’s quality. Every day the samples investigated by Cobas Integra 400 machines.

### Data analysis

The data were entered, analyzed using the computer software (Microsoft office excel). Descriptive statistics was applied in the form of number and percentages.

### Ethical approval

Ethical approval was obtained from the research committee at Faculty of Medical Technology, the University of Tripoli, and a permission letter was obtained from different medical polyclinics before going ahead with the study. Confidentiality was kept throughout the collection and processing of specimens.

## Results

### Prevalence of GDM

A bout 312 gestational diabetes patients (18%) were observed in 1700 pregnant women admitted to gynecology department in different medical polyclinics at the time of study.

**Figure 1.**
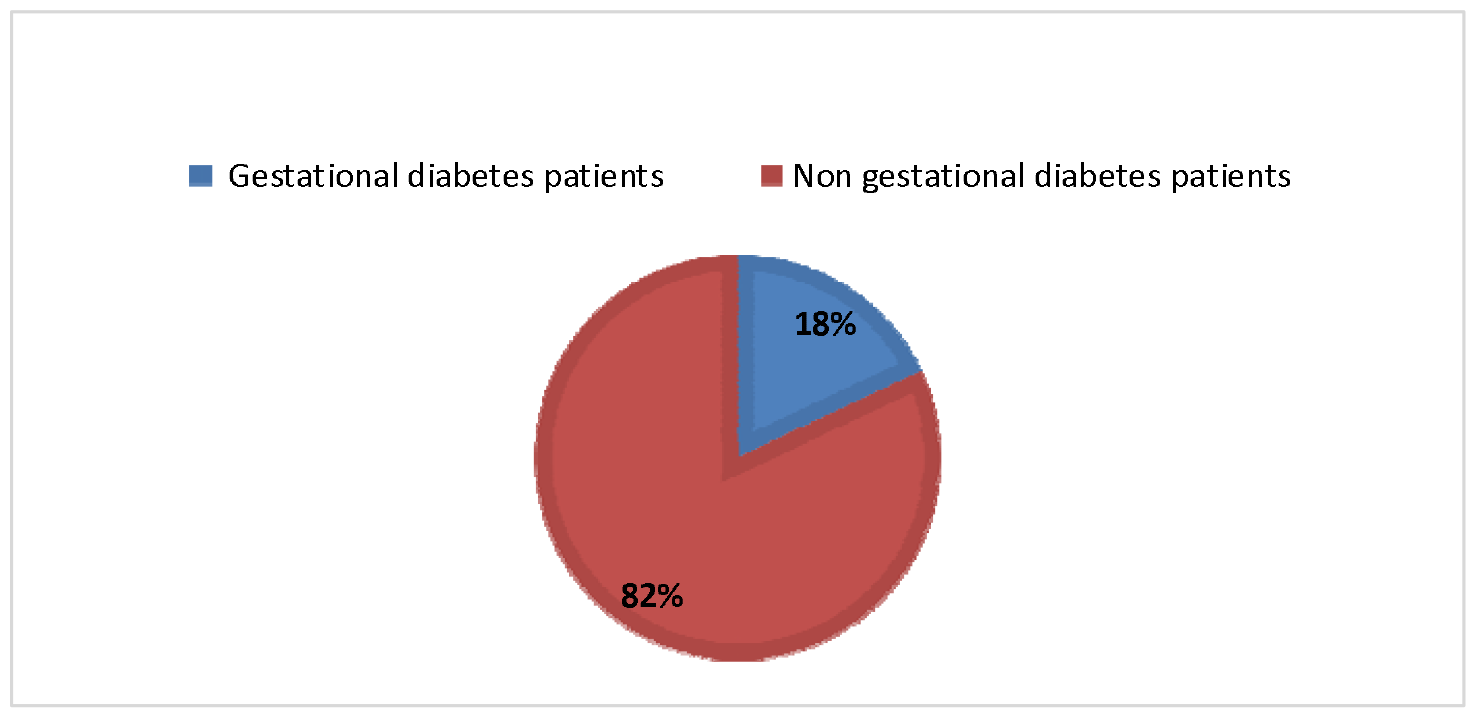
The results of total samples.

### Relation between age and GDM

GDM increased with increase of age where it raised from 23% (74 patients) in age groups 20-29 years old and then 31 % (96 patients) in age group 30-39-year-old to reach maximum 46% (142 patients) in age group 40-49 years old.

**Figure 2.**
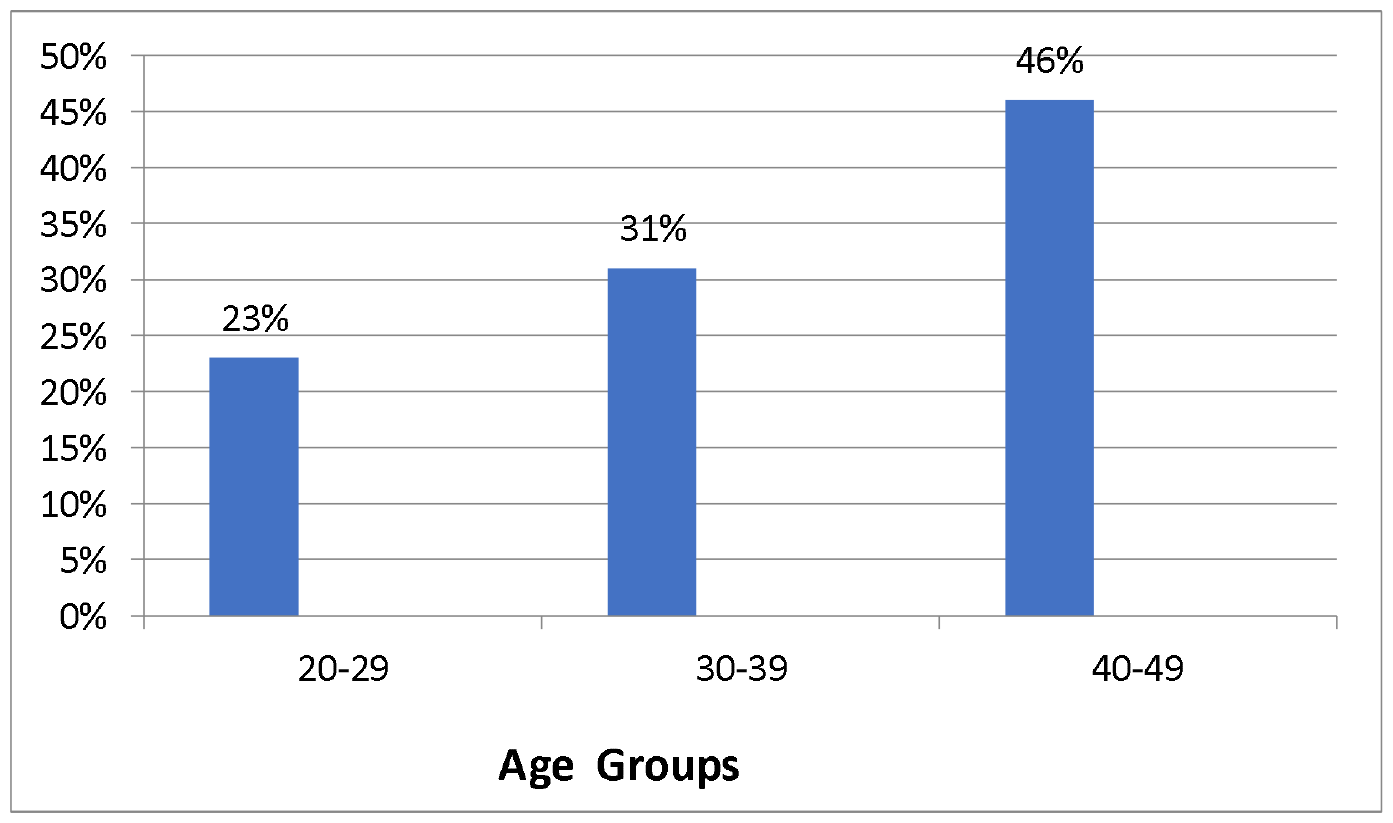
The relation between age and incidence of GDM

### Stages of pregnancy associated GDM

The prevalence of GDM in pregnant women increased with increase of the gestational age to reach maximum at the third trimester of gestation. Onset of GDM pregnant women were 3% (9 patients) during first trimester, 11% (34 patients) in second trimester and 86% (269 patients) in third trimester.

**Table1.** Prevalence of GDM during the pregnancy stages.

| Gestational trimester | Number of patients % |
| --- | --- |
| First trimester | 9 (3% ) |
| Second trimester | 34 (11% ) |
| Third trimester | 269 (86%) |

### Prevalence of anemia in GDM patients

This result indicates that 31% (96 patients) anemic pregnant women were observed in 312 gestational diabetes patients.

**Figure 3.**
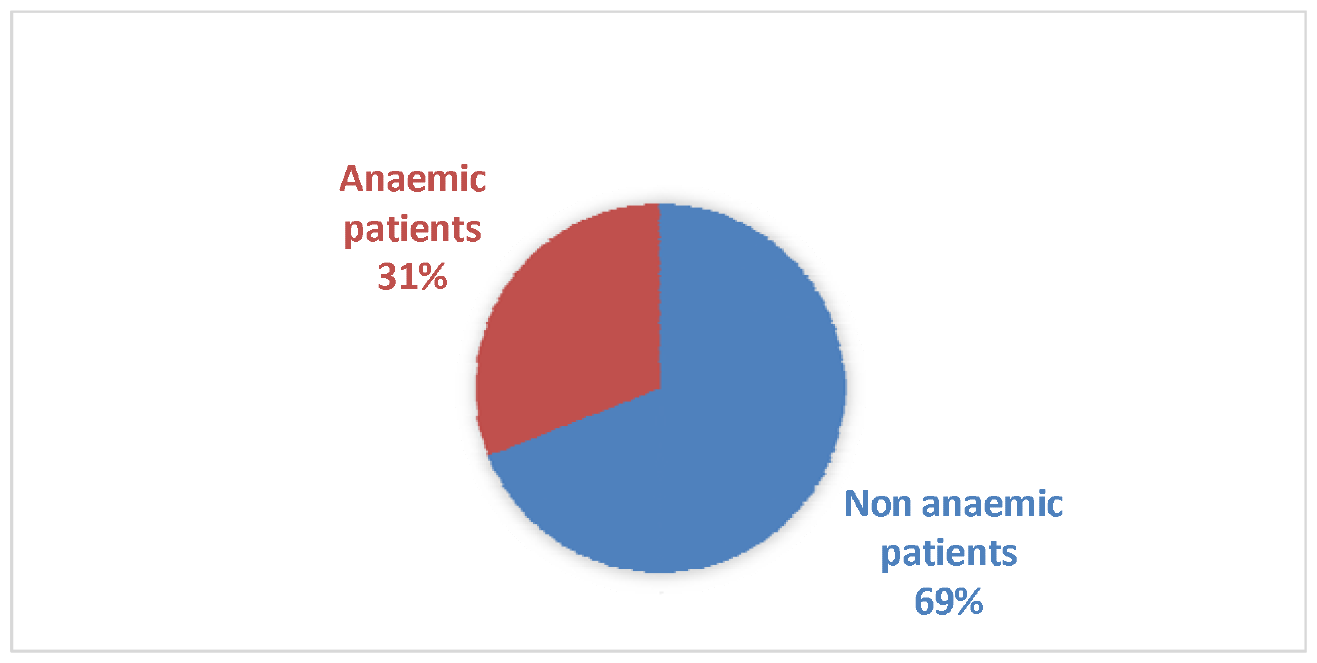
The incidence of anemia in gestational diabetes patients.

### Degree of anemia in GDM patients

Data shows that 19% (59 Patients) of patients had mild anemia and 10% (31 patients) of patients had moderate anemia and 2% (6 patients) severe anemia cases were found in this study when compared with 69% (216 patients) non anemic.

**Table 1.** The degrees of anemia in GDM patients.

| Degree of anemia | Number of patients (%) |
| --- | --- |
| Mild | 59 ( 19%) |
| Moderate | 31 (10% ) |
| Severe | 6 (2% ) |

### Relation between degree of anemia and stages of pregnancy

The prevalence of anemia was 0.3% (1 patient) during first trimester with mild anemia, 0.3% (1 patient) with moderate anemia and 0.3% (1 patient) with severe anemia. While the prevalence of anemia was 5.7% (17 patients) during second trimester with mild anemia, 4% (12 patients) with moderate anemia and 1.4% (4 patients) with severe anemia. Moreover, 13% (41 patients) in third trimester with mild anemia and 5.7% (18 patients) with moderate anemia, 2% (6 patients) with severe anemia.

**Table 2.**
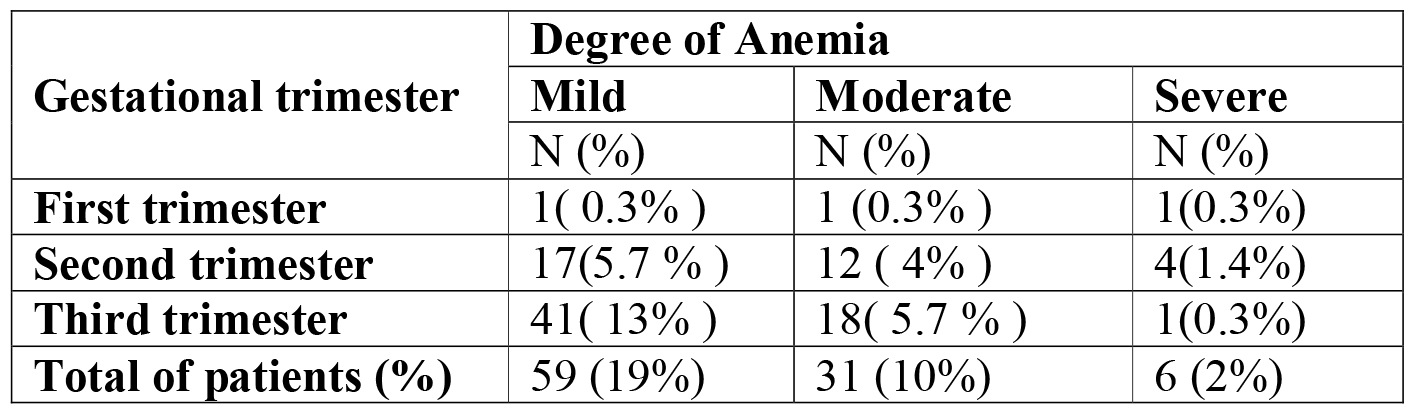
The relation between degree of anemia and stages of pregnancy.

| Gestational trimester | Degree of Anemia |  |  |
| --- | --- | --- | --- |
|  | Mild | Moderate | Severe |
|  | N (%) | N (%) | N (%) |
| First trimester | 1( 0.3% ) | 1 (0.3% ) | 1(0.3%) |
| Second trimester | 17(5.7 % ) | 12 ( 4% ) | 4(1.4%) |
| Third trimester | 41( 13% ) | 18( 5.7 % ) | 1(0.3%) |
| Total of patients (%) | 59 (19%) | 31 (10%) | 6 (2%) |

## Discussion

There are many researches explored the prevalence and cause of GDM during pregnancy worldwide [13,14]. In the present study, the prevalence of gestational diabetes disorder among pregnant women were 18% of pregnant women. This in quite similar to the observation from a previous studies conducted in Peru that reported prevalence of GDM was 16% [15]. On the contrary, the observed prevalence of this study higher compared to others studies done in Ghana 8.5% [16], Nigeria 8.3% [17], 10% in Korea [18], and 6% in the USA [19]. However, the obtained result in the current study was lower than the GDM prevalence among Bangladeshi pregnant women was 35% [20]. In present study, gestational diabetes increased with increase of age where it raised from 23% (74 patients) in age groups 20-29 years old and then 31% (96 patients) in age group 30–39-year-old to reach maximum 46% (142 patients) in age group 40-49 years old. In similar study, Singh et al., [21] recorded that gestational diabetes in pregnant women were 29.0±4.9 years, and concluded that increased maternal age is an important risk factor for the development of GDM. Also, Parnas et al., [22] reported that pregnant women with gestational diabetes were significantly older (30.7 ± 5.9 versus 28.7 ± 5.7; p = 0.001) compared with pregnant women without gestational diabetes.

GDM is frequently detected during the second and third trimesters of pregnancy [23]. A previous study had observed a trend between GDM and pregnancy trimester. The prevalence of GDM in the middle east and north Africa regions reported to be increased by 45.0%, from 8.9% in the first trimester to 12.9% in the second trimester, and by 55.0% in the third trimester (20.0%, 95% CI, 13.1–27.9%, I2, 98.8%) compared with the second trimester [24]. In our study, we observed similar results, an increase of the gestational age to reach maximum (86%) at the third trimester of gestation, compared to 11% in the second trimester.

It was hypothesized that GDM may contribute to maternal anemia and that an anti-inflammatory diet can alleviate this negative effect [25]. In the current study, 31% of the GDM patients were anemic, majority of them were with mild and moderate anemia. A recent study conducted in 2023 also reported higher incidence of moderate anemia among pregnant women with GDM compared to pregnant without GDM (40.0% vs. 11.4%, P = .029) [25]. Iron-rich foods and supplements are often consumed as part of dietary interventions for the prevention and treatment of maternal anemia. However, because of these treatments, maternal anemia is more likely to develop in GDM women with higher inflammatory statuses.

## Conclusion

The prevalence of GDM in Libya was high in pregnant women aged >39 years (46%), in their third trimester (86%). Current evidence suggests that GDM may increase a pregnant woman’s chance of developing anemia. Careful surveillance is required for these pregnancies for early detection and treatment of possible complications, in order to try to reduce maternal and neonatal morbidities. Further prospective studies investigating the link between GDM and anemia are warrant.

## Data Availability

Data available upone request

